# Using the Index of Concentration at the Extremes to Evaluate Associations of Income and Black-White Racial Segregation with HIV Outcomes Among Adults Aged ≥18 Years — United States and Puerto Rico, 2019

**DOI:** 10.1101/2023.01.25.23285022

**Authors:** Zanetta Gant, André Dailey, Xiaohong Hu, Wei Song, Linda Beer, Shacara Johnson Lyons, Damian J. Denson, Anna Satcher Johnson

## Abstract

**Objective(s):** To examine associations between Index of Concentration at the Extremes (ICE) measures for economic and racial segregation and HIV outcomes in the United States (U.S.) and Puerto Rico.

**Methods:** County-level HIV testing data from CDC’s National HIV Prevention Program Monitoring and Evaluation and census tract-level HIV diagnoses, linkage to HIV medical care, and viral suppression data from the National HIV Surveillance System were used. Three ICE measures of spatial polarization were obtained from the U.S. Census Bureau’s American Community Survey: ICEincome (income segregation), ICErace (Black-White racial segregation), and ICEincome+race (Black-White racialized economic segregation). Rate ratios (RRs) for HIV diagnoses and prevalence ratios (PRs) for HIV testing, linkage to care within 1 month of diagnosis, and viral suppression within 6 months of diagnosis were estimated with 95% confidence intervals (CIs) to examine changes across ICE quintiles using the most privileged communities (Quintile 5, Q5) as the reference group.

**Results:** PRs and RRs showed a higher likelihood of testing and adverse HIV outcomes among persons residing in Q1 (least privileged) communities compared with Q5 (most privileged) across ICE measures. For HIV testing percentages and diagnosis rates, PRs and RRs were consistently greatest for ICErace. For linkage to care and viral suppression, PRs were consistently lower for ICEincome+race.

**Conclusions:** Income, racial, and economic segregation—as measured by ICE—might contribute to poor HIV outcomes and disparities by unfairly concentrating certain groups (i.e., Black persons) in highly segregated and deprived communities that experience a lack of access to quality, affordable health care. Expanded efforts are needed to address the social/economic barriers that might impede access to HIV care among Black persons. Increased partnerships between government agencies and the private sector are needed to change policies that promote and sustain racial and income segregation.

## Introduction

HIV continues to disproportionately affect Black/African American (Black) persons in the United States (U.S). Research reveals that socioeconomic differences between races account for a substantial portion of the racial disparity in many health outcomes, including infant mortality, heart disease, and cancer (1,2). At the same time, adjusting for socioeconomic differences does not eliminate racial disparities for all health outcomes. In other words, there is an independent contribution of racial status to disparities in specific health outcomes. These residual health differences may be due to a history of racial discrimination and residential segregation, as manifestations of structural racism which has been recognized as a primary cause of health disparities (3–5).

Structural racism is defined as the “totality of ways in which societies foster [racial] discrimination, via mutually reinforcing [inequitable] systems…(e.g., in housing, education, employment, earnings, benefits, credit, media, health care, criminal justice, etc.) that in turn reinforce discriminatory beliefs, values, and distribution of resources” (6,7). Structural racism is reflected in history, culture, and interconnected institutions and includes the most influential socioecological levels at which racism may affect racial and ethnic health inequities (8). Structural mechanisms (e.g., residential segregation) do not require the actions or intent of individuals, as they are constantly reconstituting the conditions necessary to ensure their perpetuation (8–10). Even if interpersonal discrimination were eliminated, racial inequities would likely remain unchanged due to the persistence of structural mechanisms such as residential segregation (11).

Residential segregation has been a central mechanism by which racial inequality has been created and reinforced in the U.S. and has limited the socioeconomic mobility of Black persons by determining access to educational and employment opportunities (12). Black persons are more segregated than other U.S. racial/ethnic minority groups (13). Segregation, racial and economic, is a neglected but enduring legacy of racism in the U.S. and is a factor that contributes to higher rates of HIV diagnoses and poor health outcomes among Black persons. It does this by isolating Black persons from access to important resources and affecting neighborhood quality, with populations residing in lower income and relatively more isolated areas being more vulnerable (13–16). Black persons tend to reside in communities with the highest social vulnerability in the U.S. (13,17,18). Understanding the role of community-level social and structural factors—such as racial and economic segregation—is necessary to address these racial inequities.

Using methods such as the Index of Concentration at the Extremes (ICE) to explore income and racial segregation as proxies for structural racism is necessary to understand and address HIV diagnosis and care inequities that effect certain groups (19,20). ICE measures the extent to which an area’s residents are concentrated into groups at the extremes of deprivation and privilege, also referred to as spatial social polarization (21).

Assessing the role of segregation in contributing to poor health outcomes can provide information to inform interventions to increase health equity by addressing the inequitable concentration of Black persons in U.S. areas of deprivation. This paper examines associations between ICE measures for racial and economic segregation and HIV outcomes—specifically HIV testing, HIV diagnoses, linkage to HIV medical care, and viral suppression.

### Methods

Data were obtained from 3 sources: Centers for Disease Control and Prevention’s (CDC’s) National HIV Prevention Program Monitoring and Evaluation (NHM&E) and National HIV Surveillance System (NHSS), and from the U.S. Census Bureau’s American Community Survey 2015–2019 5-year estimates (ACS).

### NHM&E

County-level HIV testing data for the U.S. and Puerto Rico were submitted to CDC by 60 CDC-funded state and local health departments and 100 community-based organizations. Data included 2019 HIV testing data for adults aged _≥_18 years that were linked to the ACS indicators using the 5-year estimates for 2015–2019 at the county level.

### NHSS

Census tract-level data on HIV diagnoses, linkage to HIV medical care within 1 month of HIV diagnosis, and viral suppression within 6 months of HIV diagnosis were obtained from NHSS for adults aged _≥_18 years with HIV diagnosed during 2019 in the U.S. and Puerto Rico. Linkage to care was measured by documentation of _≥_1 CD4 or viral load (VL) tests _≤_1 month of HIV diagnosis. A VL test result of <200 copies/mL indicates HIV viral suppression. VL test results were from the tests performed within 6 months of HIV diagnosis. Forty-six jurisdictions (45 states and the District of Columbia) submitted complete CD4 and viral load results to CDC to determine linkage to HIV medical care within 1 month and viral suppression within 6 months of HIV diagnosis. Data were not included for states and associated counties that do not have laws requiring reporting of all CD4 and viral load results or that had incomplete reporting of laboratory data to CDC. Areas without laws were Idaho and New Jersey. Areas with incomplete reporting were Kansas, Kentucky, Pennsylvania, Vermont, and Puerto Rico.

Data included NHSS case data for adults aged _≥_18 years with HIV diagnosed during 2019. Cases were geocoded to the U.S. census tract level based on residential address at the time of HIV diagnosis and linked to ACS indicators using the 5-year estimates for 2015–2019.

### ACS

Three county-level and census tract-level ICE measures of spatial polarization were obtained from the 2015–2019 5-year ACS estimates, ICEincome, ICErace, and ICEincome+race.

We computed the ICE by using the following formula (21):

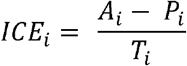

where

A_*i*_ = No. of privileged persons in county or census tract *i* (i.e., most privileged communities)

P_*i*_ = No. of deprived persons in county or census tract *i* (i.e., most deprived communities)

T_*i*_ = Total population with known information in county or census tract *i*

ICE ranges from -1, indicating 100% of the population is concentrated in the most deprived group to 1, indicating that 100% of the population is concentrated into the most privileged group. The ICE measures were categorized by quintiles, with Quintile 1 (Q1) representing the most deprived and Quintile 5 (Q5) representing the most privileged.

The 3 ICE measures were calculated as:

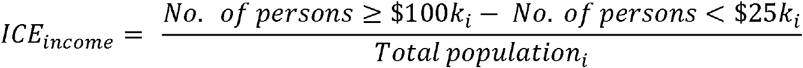

referred to as income segregation, where positive values indicate counties or census tracts with larger concentrations of persons living in households with annual incomes _≥_$100,000, and negative values indicate counties or census tracts with larger concentrations of persons living in households with annual incomes <$25,000.

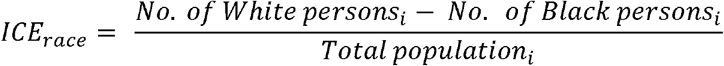

referred to as Black-White racial segregation, where positive values indicate counties or census tracts with larger concentrations of White residents, and negative values indicate counties or census tracts with larger concentrations of Black residents.

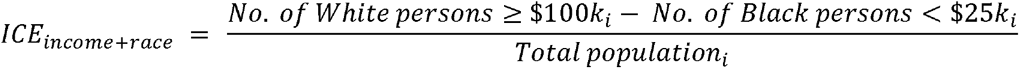

referred to as Black-White racialized economic segregation, where positive values indicate counties or census tracts with larger concentrations of White residents living in households with annual incomes _≥_$100,000, and negative values indicate counties or census tracts with larger concentrations of Black residents living in households with annual incomes <$25,000.

### Analysis

To assess the effects of income and racial segregation on the four HIV outcomes among adults aged _≥_18 years, data were analyzed to determine differences in HIV outcomes by ICE quintiles. HIV diagnosis rates were calculated per 100,000 persons. The rate ratios (RRs) for HIV diagnoses and prevalence ratios (PRs) for testing, linkage, and viral suppression were estimated with 95% confidence intervals (CIs) to examine changes in HIV outcomes across ICE quintiles; Q5 (most privileged) was the reference group. The PR and RR 95% CIs that excluded 1 were considered statistically significant. Analyses were conducted using SAS software (version 9.4; SAS Institute).

## Results

Of 1,833,877 CDC-funded HIV tests administered to adults in 2019, testing percentages were highest in Q2 for ICEincome (0.95%) and Q1 for ICErace (1.06%) and ICEincome+race (1.44%) (Table 1). Testing percentages were lowest in the most privileged (Q5) counties for all ICE measures (ICEincome =0.58%, ICErace =0.14%, and ICEincome+race =0.43%). For all ICE measures, residing in Q1 through Q4 compared to the most privileged quintile (Q5) increased the likelihood of receiving a CDC-funded HIV test. Additionally, across all 3 ICE measures, the greatest PRs (i.e., higher likelihood in Q1 compared with Q5) for HIV testing were observed for ICErace (PR = 7.50; CI = 7.39–7.63) followed by ICEincome+race (PR = 3.38; CI = 3.36–3.39).

**TABLE 1.** HIV testing among adults aged _≥_18 years, by Index of Concentration at the Extremes (ICE), 2019 — county level^a^, United States and Puerto Rico.

| HIV testing (ICE quintiles<sup>f</sup>) | ICE income<sup>b</sup> |  |  | ICE race (White-Black)<sup>c</sup> |  |  | ICE income and race (White-Black)<sup>d</sup> |  |  |
| --- | --- | --- | --- | --- | --- | --- | --- | --- | --- |
|  | Total tested | %<sup>e</sup> | PR (95% CI) | Total tested | %<sup>e</sup> | PR (95% CI) | Total tested | %<sup>e</sup> | PR (95% CI) |
| Q1 (most deprived) | 112,146 | 0.87 | 1.49 (1.48–1.50) | 1,029,830 | 1.06 | 7.50 (7.39–7.63) | 506,952 | 1.44 | 3.38 (3.36–3.39) |
| Q2 | 211,512 | 0.95 | 1.62 (1.61–1.63) | 580,517 | 0.70 | 4.92 (4.84–5.00) | 445,008 | 0.75 | 1.77 (1.76–1.77) |
| Q3 | 253,869 | 0.78 | 1.34 (1.32–1.34) | 163,175 | 0.38 | 2.69 (2.64–2.73) | 272,803 | 0.76 | 1.77 (1.77–1.78) |
| Q4 | 478,563 | 0.90 | 1.54 (1.53–1.54) | 45,004 | 0.23 | 1.61 (1.57–1.64) | 290,905 | 0.59 | 1.39 (1.38–1.40) |
| Q5 (most privileged) | 777,787 | 0.58 | Reference | 15,351 | 0.14 | Reference | 318,209 | 0.43 | Reference |

**Abbreviations:** ICE = Index of Concentration at the Extremes; PR, prevalence ratio; Q1 = quintile 1; Q2 = quintile 2; Q3 = quintile 3; Q4 = quintile 4; Q5 = quintile 5; CDC, the Centers for Disease Control and Prevention [footnotes only].
**Note.** Data obtained from CDC's National HIV Prevention Program Monitoring and Evaluation. Data were submitted by 60 CDC-funded state and local health departments and 100 community-based organizations to CDC.
<sup>a</sup>Data reflect the county of the person's residential address at the time they received an HIV test.
<sup>b</sup>ICE income = (No. of persons ≥\$100k – No. of persons <\$25k) / total population.
<sup>c</sup>ICE race = (No. of White persons – No. of Black persons) / total population.
<sup>d</sup>ICE income and race = (No. of White persons with annual income ≥\$100k – No. of Black persons with annual income <\$25k) / total population.
<sup>e</sup>Row percent.
<sup>f</sup>ICE ranges from –1.0, indicating 100% of the population is concentrated in the most deprived group, to 1.0, indicating that 100% of the population is concentrated in the most privileged group. ICE measures were categorized into quintiles based on distribution among all U.S. counties from U.S. Census Bureau's American Community Survey 2015–2019 5-year estimates. PRs with 95% CIs were calculated to examine the relative difference across quintiles with Q5 (most privileged) counties; Q5 served as the reference group. Percentages were considered significantly different if the 95% CIs of the PRs excluded 1.0.
ICE quintile cutpoints:
ICE\_income: median = –0.02; interquartile range (IQR) = –0.07 to 0.03; Q1 = –0.32 to –0.08; Q2 = –0.08 to –0.04; Q3 = –0.04 to –0.002; Q4 = –0.002 to 0.04; Q5 = 0.04 to 0.30;
ICE\_race: median = 0.81; IQR = 0.51 to 0.92; Q1 = –0.84 to 0.42; Q2 = 0.42 to 0.72; Q3 = 0.72 to 0.87; Q4 = 0.87 to 0.93; Q5 = 0.93 to 1.00;
ICE\_income\_race: median = 0.08; IQR = 0.05 to 0.11; Q1 = –0.25 to 0.04; Q2 = 0.04 to 0.07; Q3 = 0.07 to 0.09; Q4 = 0.09 to 0.12; Q5 = 0.12 to 0.27.

Among the 29,889 adults who received an HIV diagnosis in 2019, diagnosis rates were highest in Q1 for all ICE measures (ICEincome = 22.5; ICErace = 28.2; ICEincome+race = 29.9) (Table 2), and lowest in Q5 for all ICE measures (ICEincome = 5.7; ICErace = 2.8; ICEincome+race = 4.9). For all ICE measures, residing in Q1 through Q4 compared to the most privileged quintile (Q5) increased the likelihood of receiving a diagnosis of HIV infection. Additionally, across all 3 ICE measures, the greatest RRs (i.e., higher likelihood in Q1 compared with Q5) for HIV diagnosis were observed for ICErace (RR = 9.93; CI = 9.39–10.50) followed by ICEincome+race (RR = 6.06; CI = 5.82–6.32).

**TABLE 2.** Diagnoses of HIV infection, linkage to HIV medical care within 1 month, and viral suppression within 6 months of HIV diagnosis among adults aged _≥_18 years, by Index of Concentration at the Extremes (ICE) — United States and Puerto Rico (census tract level^a^), 2019.

| HIV outcomes by ICD quintile<sup>e</sup> | ICE income<sup>b</sup> |  |  | ICE race (White-Black)<sup>c</sup> |  |  | ICE income and race (White-Black)<sup>d</sup> |  |  |
| --- | --- | --- | --- | --- | --- | --- | --- | --- | --- |
|  | Total | Rate | RR (95% CI) | Total | Rate | RR (95% CI) | Total | Rate | RR (95% CI) |
| **HIV diagnosis** |  |  |  |  |  |  |  |  |  |
| Q1 (most deprived) | 8,841 | 22.5 | 3.94 (3.78–4.10) | 13,038 | 28.2 | 9.93 (9.39–10.50) | 11,776 | 29.9 | 6.06 (5.82–6.32) |
| Q2 | 6,349 | 13.3 | 2.33 (2.23–2.43) | 7,612 | 13.8 | 4.85 (4.58–5.14) | 7,273 | 12.2 | 2.47 (2.36–2.57) |
| Q3 | 5,247 | 10.2 | 1.79 (1.71–1.87) | 4,504 | 8.2 | 2.90 (2.73–3.08) | 3,245 | 7.5 | 1.53 (1.45–1.60) |
| Q4 | 4,772 | 8.0 | 1.39 (1.33–1.46) | 2,404 | 4.8 | 1.71 (1.60–1.82) | 3,223 | 6.0 | 1.21 (1.15–1.28) |
| Q5 (most privileged) | 3,162 | 5.7 | Reference | 1,361 | 2.8 | Reference | 2,852 | 4.9 | Reference |
| Missing | 1,518 | — | — | 969 | — | — | 1,518 | — | — |
| **Linkage to care<sup>f</sup>** |  |  |  |  |  |  |  |  |  |
| Q1 (most deprived) | 6,321 | 79.6 | 0.94 (0.92–0.96) | 9,495 | 80.0 | 0.96 (0.93–0.98) | 8,480 | 79.3 | 0.93 (0.91–0.95) |
| Q2 | 4,759 | 80.5 | 0.95 (0.93–0.97) | 5,916 | 82.8 | 0.99 (0.96–1.02) | 5,603 | 82.1 | 0.96 (0.94–0.98) |
| Q3 | 4,012 | 82.4 | 0.97 (0.95–0.99) | 3,483 | 83.6 | 1.00 (0.97–1.03) | 2,514 | 83.5 | 0.98 (0.96–1.00) |
| Q4 | 3,719 | 84.0 | 0.99 (0.97–1.01) | 1,790 | 83.4 | 1.00 (0.97–1.03) | 2,456 | 83.6 | 0.98 (0.96–1.00) |
| Q5 (most privileged) | 2,439 | 84.5 | Reference | 950 | 83.6 | Reference | 2,198 | 85.3 | Reference |
| Missing | 1,094 | 81.7 | — | 711 | 77.6 | — | 1,094 | 81.7 | — |
| **Viral suppression<sup>f</sup>** |  |  |  |  |  |  |  |  |  |
| Q1 (most deprived) | 5,276 | 66.4 | 0.91 (0.89–0.94) | 8,047 | 67.8 | 0.93 (0.89–0.96) | 7,135 | 66.7 | 0.89 (0.86–0.91) |
| Q2 | 4,056 | 68.6 | 0.94 (0.92–0.97) | 5,046 | 70.6 | 0.96 (0.93–1.00) | 4,805 | 70.5 | 0.94 (0.91–0.96) |
| Q3 | 3,455 | 70.9 | 0.97 (0.95–1.00) | 3,034 | 72.8 | 1.00 (0.96–1.04) | 2,172 | 72.2 | 0.96 (0.93–0.99) |
| Q4 | 3,291 | 74.3 | 1.02 (0.99–1.05) | 1,565 | 73.0 | 1.00 (0.95–1.04) | 2,124 | 72.3 | 0.96 (0.93–0.99) |
| Q5 (most privileged) | 2,100 | 72.8 | Reference | 832 | 73.2 | Reference | 1,942 | 75.3 | Reference |
| Missing | 946 | 70.6 | — | 600 | 65.5 | — | 946 | 70.6 | — |

**Abbreviations:** CI = confidence interval; ICE = Index of Concentration at the Extremes; PR = prevalence ratio; Q1 = quintile 1; Q2 = quintile 2; Q3 = quintile 3; Q4 = quintile 4; Q5 = quintile 5; RR = rate ratio; VL = viral load; CDC, the Centers for Disease Control and Prevention [footnotes only].
**Note.** Data obtained from the National HIV Surveillance System. Data reflect the census tract of the person's residential address at the time of receipt of an HIV diagnosis.
<sup>a</sup>Data reflect the census tract of the person's residential address at the time of receipt of an HIV diagnosis. Rates = cases per 100,000 population
<sup>b</sup>ICE income = (No. of persons ≥ \$100k – No. of persons $<$ \$25k) / total population.
<sup>c</sup>ICE race = (No. of White persons – No. of Black persons) / total population.
<sup>d</sup>ICE income and race = (No. of White persons with annual income ≥ \$100k – No. of Black persons with annual income $<$ \$25k) / total population.
<sup>e</sup>ICE ranges from –1.0, indicating 100% of the population is concentrated in the most deprived group, to 1.0, indicating that 100% of the population is concentrated in the most privileged group. ICE measures were categorized into quintiles based on distribution among all U.S. census tracts from U.S. Census Bureau's American Community Survey 2015–2019 5-year estimates. RRs and PRs with 95% CIs were calculated to examine the relative difference across quintiles with Q5 (most privileged) census tracts; Q5 served as the reference group. Rates and percentages were considered significantly different if the 95% CIs of the RRs or PRs excluded 1.0.
ICE quintile cutpoints:
ICE\_income: median = 0.03; interquartile range (IQR) = –0.07 to 0.13; Q1 = –1.00 to –0.09; Q2 = –0.09 to –0.01; Q3 = –0.01 to 0.06; Q4 = 0.06 to 0.16; Q5 = 0.16 to 1.00;
ICE\_race: median = 0.64; IQR = 0.23 to 0.86; Q1 = –1.00 to 0.11; Q2 = 0.11 to 0.51; Q3 = 0.51 to 0.75; Q4 = 0.75 to 0.89; Q5 = 0.89 to 1.00;
ICE\_income\_race: median = 0.08; IQR = 0.01 to 0.15; Q1 = –1.00 to 0; Q2 = 0 to 0.06; Q3 = 0.06 to 0.10; Q4 = 0.10 to 0.16; Q5 = 0.16 to 1.00.
<sup>f</sup>Linkage to care and viral suppression were assessed using data from 45 areas with complete reporting of lab data to CDC as of December 2021.
Linkage to HIV medical care was measured by documentation of ≥ 1 CD4 or VL tests ≤ 1 month after HIV diagnosis. A VL test result of $<$ 200 copies/mL indicates HIV viral suppression. VL test results are within 6 months of diagnosis of HIV infection during 2019. Data not provided for states and associated census tracts that do not have laws requiring reporting of all CD4 and viral loads or that have incomplete reporting of laboratory data to CDC Areas without laws: Idaho and New Jersey. Areas with incomplete reporting: Kansas, Kentucky, Pennsylvania, Vermont, and Puerto Rico.

For linkage to HIV medical care within 1 month of diagnosis in 2019, the lowest percentages were in Q1 for all ICE measures ICEincome = 79.6%; ICErace = 80.0%; ICEincome+race = 79.3%, and highest in Q5 for all ICE measures (ICEincome = 84.5%; ICErace = 83.6%; ICEincome+race = 85.3%) (Table 2). For viral suppression within 6 months of diagnosis in 2019, the lowest percentages were lowest in Q1 for all ICE measures (ICEincome = 66.4%; ICErace = 67.8%; ICEincome+race = 66.7%) and highest in Q4 for ICEincome (74.3%) and Q5 for ICErace = 73.2%) and ICEincome+race (75.3%) (Table 2). For all ICE measures where statistically significant differences were found, residing in Q1 through Q4 compared to Q5 decreased the likelihood of being linked to HIV medical care within 1 month of diagnosis or to have viral suppression within 6 months of diagnosis. Across all 3 ICE measures for linkage to care and viral suppression, the smallest PRs (i.e., lower likelihood in Q1 compared with Q5) were observed for ICEincome+race (linkage, PR = 0.93; CI = 0.91–0.95; viral suppression, PR = 0.89; CI = 0.86–0.91) followed by ICEincome (linkage, PR = 0.94; CI = 0.92–0.96; viral suppression, PR = 0.91; CI = 0.89– 0.94).

## Discussion

This analysis is the first large-scale county-level and census tract-level analysis to utilize ICE to assess the relationship between racial and economic segregation on HIV outcomes across the U.S and Puerto Rico. This analysis found that adults who resided in the most privileged communities (Q5) have substantially better HIV outcomes than adults in the most deprived communities (Q1). For income, Black-White racial segregation, and Black-White racialized economic segregation, higher HIV testing percentages and diagnosis rates and lower linkage to HIV medical care and viral suppression percentages were observed in the most deprived compared with the most privileged communities. The highest PRs for HIV testing percentages and RRs for diagnosis rates were observed for ICErace, and lowest PRs for linkage to care and viral suppression were observed for ICEincome+race.

Our findings of higher percentages of HIV testing in more deprived communities can be explained by CDC-funded testing efforts being focused on high-priority populations, and this testing might partially explain the higher HIV diagnosis rates in these communities. However, SDOH factors shaped by income, education, wealth, and childhood and neighborhood socioeconomic conditions, which vary systematically by race/ethnicity groups, might also explain higher diagnosis rates as well as our finding of lower percentages of linkage to care and viral suppression in these communities (14,22,23). These findings suggest spatial social polarization, as demonstrated by the ICE measures, might contribute to poor HIV outcomes and disparities for Black adults by segregating them in more deprived communities (14).

For HIV testing percentages and diagnosis rates, PRs and RRs were consistently greatest for ICErace, where communities with the highest concentrations of Black residents had higher testing percentages and diagnosis rates than communities with the highest concentrations of White residents. For linkage to care and viral suppression, PRs were consistently lower for ICEincome+race, where communities with the highest concentrations of Black residents living in households with annual incomes <$25,000 had linkage and viral suppression percentages that were lower than those of communities with the highest concentrations of White residents living in households with annual incomes _≥_$100,000. This is consistent with previous research that examined racial residential segregation (24). Residential segregation remains pervasive and may influence health by concentrating poverty, environmental pollutants, infectious agents, and other adverse conditions (12, 25). For instance, Morello-Frosch and Jesdale (26) found that segregation increased the risk of cancer related to air pollution. Studies using multilevel modeling that simultaneously accounts for individual and structural factors also find associations between segregation and illness (27,28). Our findings suggest that Black-White racial segregation and Black-White racialized economic segregation contribute to adverse health outcomes more than income segregation alone, in other words, Black-White racial segregation alone or in combination with economic segregation plays an important role in the production of inequitable and adverse health outcomes (e.g., lack of access to quality, affordable health care) by unfairly concentrating Black persons in highly segregated, deprived communities (14).

To our knowledge, this is the first time ICE has been used to analyze county-level and census tract-level data for HIV outcomes. The use of this novel measure provides evidence to support our hypothesis that the worst HIV outcomes (i.e., diagnosis, linkage to care, viral suppression) occur in the most deprived communities. What’s also new and important is that it adds to the literature by quantifying the negative effects that structural racism (as measured by income, racial, and racialized economic segregation) has on HIV care and treatment outcomes when there is more Black-White racial segregation. These results can be used to inform policy and programmatic efforts that support investments in these communities and equitable redistribution of resources that improves the health of all persons.

This work further suggests that more action and innovative strategies are needed to achieve HIV diagnosis and treatment equity when there is increased Black-White segregation. For example, an innovative strategy might include use of implementation science from the Ryan White Special Projects of National Significance Program, which supports the development of pioneering HIV care and treatment models to quickly respond to emerging needs of persons with HIV (29). Accelerated implementation of HIV testing strategies that include rapid linkage to care and treatment is needed to identify persons with infection to increase viral suppression or linkage to prevention efforts (30). For example, the District of Columbia’s Red Carpet Entry Program is a structural-level intervention that used an improved, redesigned referral network to link 70% of persons with diagnosed HIV to care within 72 hours (31). Expanded efforts should continue to address access to health care and social and economic barriers that might impede access (30). In addition to efforts to ameliorate consequences of segregation, increased partnerships between government agencies and private sector are needed to change policies that promote and sustain segregation (32). The U.S. Department of Housing and Urban Development (HUD) recently reinstated the Fair Housing Act’s Affirmatively Furthering Fair Housing (AFFH) requirement, which requires HUD and its funding recipients to address segregation and foster inclusive communities (33).

Our analysis had several limitations. First, CDC-funded HIV testing efforts focus on high-priority populations who might reside in disadvantaged communities and might not represent national HIV testing patterns. Second, HIV diagnoses data might not be representative of all persons with HIV because not all persons with HIV have been tested or tested at a time when the infection could be detected and diagnosed. Third, linkage to care and viral suppression data were limited to 45 jurisdictions with complete reporting of laboratory data to CDC. Since CD4 and VL test results reported to HIV surveillance programs were needed to monitor the outcomes, not having these tests done or reported may prevent representation for all the outcomes in jurisdictions and monitoring of outcomes. Data on CD4 and VL test results during the follow □ up period may be delayed or missing for people who may have migrated to another jurisdiction (after HIV diagnosis) that did not report complete test results to CDC. Finally, NHSS data were limited to people whose residential addresses were complete and could be geocoded (∼85.4%) and might not reflect the entire adult population with diagnosed HIV in those census tracts.

## Conclusion

Understanding the impacts of structural racism on HIV diagnosis and care disparities, particularly among persons residing in the most deprived, highly segregated communities, can aid HIV prevention efforts and guide public health strategies and the equitable resource allocation needed to provide greater and better access to HIV care and other resources. In addition to addressing negative effects of segregation on affected communities, discontinuing policies that promote and sustain segregation will likely contribute to reducing HIV transmission and achieving health equity in the U.S. If we do not address structural racism, health disparities will continue to persist.

## Data Availability

All data produced in the present study are available in limited capacity due to the sensitive nature of the data.

## Acknowledgments

We would like to acknowledge the <u>Public Health Disparities Geocoding Project Training 2.0</u> team, based at the Harvard T.H. Chan School of Public Health (Boston, MA) for their training about the ICE and other area-based social metrics for health equity monitoring and research: Nancy Krieger (PI), Jarvis Chen, Pam Waterman, Christian Testa, Dena Javadi, Enjoli Hall, Justin Morgan, Tamara Rushovich, Sudipta Saha.

Publication of this article was made possible with the contributions of the local, state and territorial health departments and the surveillance programs and community-based organizations that provided data to CDC. The authors thank CDC colleagues for their review of and feedback on this article.

## References

1. Committee on Capitalizing on Social Science and Behavioral Research to Improve the Public’s Health, Division of Health Promotion and Disease Prevention, Institute of Medicine*. Promoting health: Intervention strategies from social and behavioral research. American Journal of Health Promotion. 2001 Jan;15(3):149–66.

2. Sondik EJ, Huang DT, Klein RJ, Satcher D. Progress toward the healthy people 2010 goals and objectives. Annual Review of Public Health. 2010 Apr 21;31:271–81.

3. Chambers BD, Baer RJ, McLemore MR, Jelliffe-Pawlowski LL. Using index of concentration at the extremes as indicators of structural racism to evaluate the association with preterm birth and infant mortality—California, 2011–2012. Journal of Urban Health. 2019 Apr;96(2):159–70.

4. Bailey ZD, Krieger N, Agénor M, Graves J, Linos N, Bassett MT. Structural racism and health inequities in the USA: evidence and interventions. The Lancet. 2017;389(10077):1453–63.

5. Gee GC, Ford CL. Structural racism and health inequities: Old issues, New Directions1. Du Bois Review: Social Science Research on Race. 2011;8(1):115–32.

6. Krieger N. Structural racism, health inequities, and the two-edged sword of data: structural problems require structural solutions. Frontiers in Public Health. 2021 Apr 15;9:655447.

7. Powell JA. Structural racism: building upon the insights of John Calmore. North Carolina Law Review. 2007; 86:791–816

8. Bailey ZD, Feldman JM, Bassett MT. How structural racism works—racist policies as a root cause of US racial health inequities. New England Journal of Medicine. 2021 Feb 25;384(8):768–73.

9. Bonilla-Silva E. Rethinking racism: Toward a structural interpretation. American Sociological Review. 1997 Jun 1:465–80..

10. Link BG, Phelan J. Social conditions as fundamental causes of disease. Journal of Health and Social Behavior. 1995 Jan 1:80–94.

11. Jones CP. Levels of racism: a theoretic framework and a gardener’s tale. American Journal of Public Health. 2000 Aug;90(8):1212.

12. Williams DR, Collins C. Racial residential segregation: a fundamental cause of racial disparities in health. Public Health Reports. 2016 Nov 30.

13. Williams DR, Jackson PB. Social sources of racial disparities in health. Health Affairs. 2005 Mar;24(2):325–34.

14. Ibragimov U, Beane S, Adimora AA, Friedman SR, Williams L, Tempalski B, Stall R, Wingood G, Hall HI, Johnson AS, Cooper HL. Relationship of racial residential segregation to newly diagnosed cases of HIV among Black heterosexuals in US metropolitan areas, 2008–2015. Journal of Urban Health. 2019 Dec;96(6):856–67.

15. Centers for Disease Control and Prevention. Social determinants of health among adults with diagnosed HIV infection, 2019. HIV Surveillance Supplemental Report 2022;27(No. 2). http://www.cdc.gov/hiv/library/reports/hiv-surveillance.html. Published March 2022. Accessed Jan 3, 2023.

16. Fennie KP, Lutfi K, Maddox LM, Lieb S, Trepka MJ. Influence of residential segregation on survival after AIDS diagnosis among non-Hispanic blacks. Annals of Epidemiology. 2015 Feb 1;25(2):113–9.

17. Braveman PA, Egerter SA, Mockenhaupt RE. Broadening the focus: the need to address the social determinants of health. American Journal of Preventive Medicine. 2011 Jan 1;40(1):S4–18.

18. Wright JE, Merritt CC. Social equity and COVIDL19: The case of African Americans. Public Administration Review. 2020 Sep;80(5):820–6.

19. CDC. Racism and health: racism is a serious threat to the public’s health. CDC; 2021. Accessed Jan 3, 2023. https://www.cdc.gov/minorityhealth/racism-disparities/index.html

20. National Institutes of Health. Structural racism and discrimination. U.S. Department of Health and Human Services, National Institutes of Health; 2022. Accessed Jan 3, 2023. https://www.nimhd.nih.gov/resources/understanding-health-disparities/srd.html

21. Krieger N, Waterman PD, Spasojevic J, Li W, Maduro G, Van Wye G. Public health monitoring of privilege and deprivation with the index of concentration at the extremes. American Journal of Public Health. 2016 Feb;106(2):256–63.

22. Williams W, Krueger A, Wang G, Patel D, Belcher L. The contribution of HIV testing funded by the Centers for Disease Control and Prevention to HIV diagnoses in the United States, 2010– 2017. Journal of Community Health. 2021 Aug;46(4):832–41.

23. Delaney KP, DiNenno EA. HIV testing strategies for health departments to end the epidemic in the US. American Journal of Preventive Medicine. 2021 Nov 1;61(5):S6–15.

24. Kramer MR, Hogue CR. Is segregation bad for your health?. Epidemiologic Reviews. 2009 Nov 1;31(1):178–94.

25. Gee GC, Payne-Sturges DC. Environmental health disparities: a framework integrating psychosocial and environmental concepts. Environmental Health Perspectives. 2004 Dec;112(17):1645–53.

26. Morello-Frosch R, Jesdale BM. Separate and unequal: residential segregation and estimated cancer risks associated with ambient air toxics in US metropolitan areas. Environmental Health Perspectives. 2006 Mar;114(3):386–93.

27. Bell JF, Zimmerman FJ, Almgren GR, Mayer JD, Huebner CE. Birth outcomes among urban African-American women: a multilevel analysis of the role of racial residential segregation. Social Science & Medicine. 2006 Dec 1;63(12):3030–45.

28. Subramanian SV, Acevedo-Garcia D, Osypuk TL. Racial residential segregation and geographic heterogeneity in black/white disparity in poor self-rated health in the US: a multilevel statistical analysis. Social Science & Medicine. 2005 Apr 1;60(8):1667–79.

29. Health Resources and Services Administration. HRSA Ryan White HIV/AIDS Program: Part F: Special Projects of National Significance (SPNS) program. Health Resources and Services Administration; 2022. Accessed Jan 3, 2023. https://ryanwhite.hrsa.gov/about/parts-and-initiatives/part-f-spns

30. Harris NS, Johnson AS, Huang YL, Kern D, Fulton P, Smith DK, Valleroy LA, Hall HI. Vital signs: status of human immunodeficiency virus testing, viral suppression, and HIV preexposure prophylaxis—United States, 2013–2018. Morbidity and Mortality Weekly Report. 2019 Dec 6;68(48):1117.

31. Olejemeh C, Denson A, Frison L, Freehill G, Pappas G. Treatment is prevention: enhanced entry into HIV care: the Red Carpet strategy in the District of Columbia, USA. Abstract presented at the XIX International AIDS Conference 2022; Washington, DC.

32. Rouse C, Bernstein J, Knudsen H, Zhang J. Exclusionary zoning: Its effect on racial discrimination in the housing market. The White House. 2021. https://www.whitehouse.gov/cea/written-materials/2021/06/17/exclusionary-zoning-its-effect-on-racial-discrimination-in-the-housing-market/

33. U.S. Department of Housing and Urban Development. Affirmatively furthering fair housing (AFFH). Accessed Jan 3, 2023. https://www.hud.gov/AFFH

